# A Framework For Reconstruction And Analysis Of Extended Pedigrees From Large-Scale Genealogy And Archive Records

**DOI:** 10.64898/2026.08.21.26360249

**Authors:** Daphne van Oosten, Paul Beele, Bi-nan Wang, Sophie Josephine Plasmans, Nienke Wolthuis, Kyara van den Berg, Maaike Paulina Theodora Blom, Myrte Meyjes, Nina Dianne van der Schoot, Hermieneke Vergunst-Bosch, Aimée Rosanne Kok, Lucas Johannes van de Ven, Michael A. van Es, Leonard Hendrik van den Berg, Jan Herman Veldink, Wouter van Rheenen

**Affiliations:** Department of Neurology, UMC Utrecht Brain Center, University Medical Center Utrecht, Utrecht, 3584 CX, The Netherlands

## Abstract

Genetic analyses in pedigrees remain a powerful approach to identify rare disease-causing variants. Reconstructing large enough pedigrees to yield sufficient power, however, still faces challenges.

Clinical family histories are often incomplete, especially in late-onset diseases caused by genetic variants with incomplete penetrance. Amyotrophic lateral sclerosis (ALS) is such a disease with a disproportionately large contribution from rare variants with high but variable penetrance. Here, we present an open-source pipeline to reconstruct extended pedigrees from large-scale archive data dating back to ∼1800, which we applied to a cohort of Dutch ALS patients carrying the *C9orf72* hexanucleotide repeat expansion (HRE). Among 238 *C9orf72* HRE carriers, 91 (38%) were members of an extended pedigree. Of these, 48 (53%) were clinically classified as sporadic ALS. In total, we identified 39 extended pedigrees including affected relatives up to the eighth degree of relatedness. Our approach increased the number of identified relationships by 2.5-fold compared with clinically obtained family histories, that typically missed relationships beyond the third degree. Genome-wide IBD analysis in pedigrees with ≥7 meioses fine-mapped the *C9orf72* HRE to shared IBD segments spanning 10.2 - 127.0 centimorgans in 97% of pedigrees. This combined genealogical–genetic approach for large-scale reconstruction of extended pedigrees supports the reclassification of apparently sporadic cases, facilitates the discovery of new disease-causing variants in ALS, and is applicable to any disease with comparable genetic architecture.

## Introduction

Family-based studies remain a powerful tool in gene discovery research and the clinical genetics practice.^1–3^ Whereas the rapid development in sequencing techniques provides major advances in profiling genetic variation, reconstructing large enough pedigrees to yield sufficient power still faces challenges. In complex and genetically heterogeneous traits, clinically obtained family histories are often incomplete or inaccurate.^4^ Reduced penetrance^5^ or undiagnosed disease in previous generations for late-onset diseases will lead to underestimation of disease occurrence among relatives.^6^ Finally, the pervasive pleiotropy among disease genes can further obscure any familial aggregation of diseases. Amyotrophic lateral sclerosis (ALS) provides a showcase example of a disease affected by all these factors.

ALS is a fatal neurodegenerative disease with a disproportionately large contribution from rare variants with high but variable penetrance.^7,8^ With a median age at onset of 65 years, the lifetime risk of ALS is 0.3% in the general population, and the heritability is estimated around 50%.^9^ Although a consensus on the definition of a positive family history is lacking,^10^ it is often stated that approximately 10–15% of patients report a family history of ALS and/or frontotemporal dementia (familial ALS, fALS), whereas the remainder are classified as sporadic (sALS).^7,11^ The hexanucleotide repeat expansion (HRE) in *C9orf72*, which is derived from a single founder, is the most common genetic cause of ALS in patients with European ancestries.^8,12,13^ Its occurrence in both familial and sporadic ALS illustrates that the dichotomy between these entities is largely artificial. There is therefore a clear need to identify genetic causes and risk factors for ALS in both unexplained familial ALS and apparently sporadic patients. The emergence of effective gene-targeted therapies further underscores the need for additional gene discovery studies.

Adopting a family-based design, pedigrees can be reconstructed by leveraging personal records in historical archives. In the Netherlands, the Dutch Center for Family History (CBG) holds all records of the Dutch civil registration introduced by Napoleon Bonaparte, including birth and death records dating back to ∼1800.^14,15^ These data can be used to identify shared ancestors between (distantly) related ALS patients and thus reconstruct their pedigree. Following the digitalization of these archives, high-throughput queries of the records allow for large-scale reconstruction of pedigrees. This enables subsequent mapping of disease loci through genome-wide identification of identity-by- descent (IBD) segments shared between distantly related patients^16^ without the need for phenotypic information on their ancestors. As a result, this approach overcomes the limitations of family-based studies based on a clinical patient-reported family history alone.

Here, we present a framework for the large-scale reconstruction of extended pedigrees that integrates genealogical data for all patients dating back to ∼1800 with relationships detected from genetic data. We demonstrate that this approach substantially improves the detection of distant familial relationships compared with clinical family history assessment and enables IBD-based identification of pathogenic variants within extended families. In addition, we present an open- source toolkit, Mangrove, for processing ancestor tables from archives and automated identification of common founders, enabling interactive visualization of pedigrees and their metadata.

## Methods

### Ethical Approval

All participants provided written informed consent for genetic testing, use of clinical data, and retrieval of genealogical records. This study was approved by the Biobank Research Ethics Committee of the University Medical Center Utrecht.

### Study Design and Participants

This retrospective cohort study included patients diagnosed with ALS at the ALS Center in the Netherlands between 2000 and 2020. All patients were confirmed to carry the *C9orf72* HRE using repeat-primed PCR.

### Data Collection

We collected clinical and genealogical records for 238 ALS patients with a *C9orf72* HRE. Age at onset was defined as the age at first reported motor symptoms, and survival as the time from symptom onset to death or >24 hours of non-invasive ventilation. We obtained ancestor tables from the Dutch National Registry of Deceased Persons (NRO), which is managed by the Dutch Center for Family History (CBG). Due to privacy concerns, birth records in the Netherlands only become publicly available 100 years after the birth. The NRO contains the personal records of individuals who died after 1939 and were registered as residents of a Dutch municipality at the time of their death.

Through the parental information in these personal records, this archive is linked to records in the Dutch civil registration, introduced by Napoleon Bonaparte. Each ancestor table includes the personal records of an individual’s ancestors, going back at least 3 generations (up to the great- grandparents). Personal records include full first and middle names, full last name (for female ancestors: maiden name), date of birth, place of birth, and date of death. In specific cases, we obtained additional data from the publicly accessible genealogy database hosted by the CBG. Throughout this report, we will refer to the initial NRO-based ancestor tables as ancestor table data, and to the additional data obtained from the public archives as public archive data.

### Pedigree Reconstruction

To process the ancestor table data and reconstruct pedigrees, we developed a semi-automated pipeline, Mangrove (Figure 1, https://github.com/dvanoosten/mangrove). First, data are cleaned and filtered; names, dates of birth, and dates of death are standardized, and initials (first 3 letters of first name, first letter of each middle name) are extracted from the first names. Places of birth are geocoded, i.e. recoded to GPS coordinates. Furthermore, a sex check is performed, and records for which either the first or last name is missing are filtered out, because this is the minimum information needed for matching.

**Figure 1:**
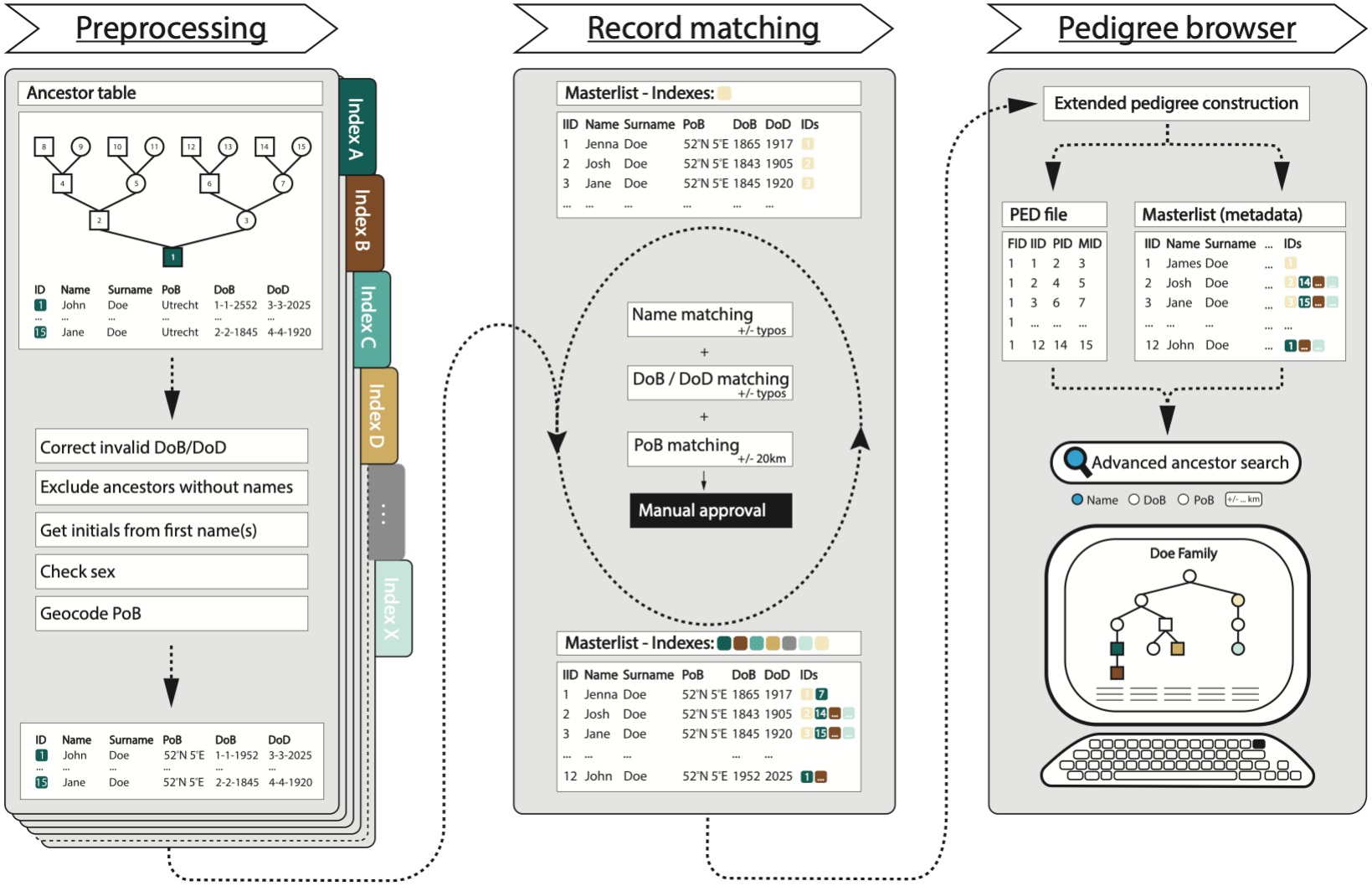
Reconstruction of ALS extended pedigrees from archive data. Workflow of genealogical data to extended pedigrees.

Next, shared ancestors are detected by cross-referencing personal data of all individuals in each ancestor table. Ancestor records are merged to create extended pedigrees when individuals are matched on sex, first name initials, last name, and, if available, date and place of birth. To account for human error, such as typos in data entry, or changing municipality borders, we allow a Damerau- Levenshtein distance <2 for the last name and date of birth, and a geographical distance <20 km for the place of birth. Matching records are manually reviewed for accuracy before they are merged to represent a common ancestor. Based on shared ancestors, Mangrove then constructs extended pedigrees and computes pairwise relationships between probands.

Finally, the data can be viewed and queried through a graphical user interface (Figure S1). One can search for all ancestors in the database using various filters, such as name or place of birth (also shown on an interactive map; Figure S1A). The pedigrees can also be accessed based on pedigree or individual IDs (Figure S1B).

### Genetic Relatedness

To assess the accuracy of reconstructed pedigrees from archive records, we used previously generated genetic data from Project MinE.^17–20^ We used two types of sequencing data: for 184 samples, whole-genome sequencing (WGS) data was available (https://projectmine.com/),^18–20^ whereas for the other 54 samples, SNP array data was imputed. Both datasets underwent QC as previously described,^18^ with the addition that the WGS data was lifted to GRCh37 (which is preferable for GRAPE^21^). Next, the two sets were merged, including only variants that were genotyped or imputed in both datasets. The data was then filtered to a minor allele frequency of 0.01 and a missingness rate of 0.1, leaving 6.3M variants. A relatedness check with KING^22^ confirmed that there were no duplicate samples.

We inferred genetic relatedness using IBD-based methods, which are shown to be reliable up to the seventh degree of relatedness.^21,23^ The GRAPE tool was applied to the dataset, using the ibis-king pipeline,^21,22,24^ with the following parameters: minimum IBD segment length set to 6.5 cM (--ibis-seg- len), a minimum of 475 SNPs per segment (--ibis-min-snp), zero-segment count threshold of 0.4 (-- zero-seg-count), zero-segment length threshold of 5.0 cM (--zero-seg-len), and an ERSA significance level of 0.01 (--alpha). A previously established definition for the degree of relatedness was followed.^23^ Importantly, GRAPE uses a slightly different definition for the relationship degree, where, for instance, full siblings and half siblings are not classified as 1^st^ and 2^nd^ degree relatives, respectively, but both as 2^nd^ degree.^21,25^ To account for this, the relationship degree was adjusted for pairs where the shared genome proportion reported by GRAPE was higher than expected based on the estimated degree.

For genetically inferred relationships not found in the ancestor table data, we retrieved more distant ancestors from the publicly accessible CBG genealogy database: WieWasWie. These records were then added to the individual’s ancestor table and imported into the dataset.

### Clinical Family History Reassessment

To compare the power of the archive-based approach with the clinical assessment of family history, we re-evaluated the clinical records of index patients across all extended pedigrees. For each patient, family history was assessed (number of affected relatives and the degree of relatedness to each affected relative) and subsequently classified as ‘familial’ or ‘sporadic’. For this, we adopted the common clinical definition of fALS; at least one first- or second-degree relative affected by ALS or FTD.^10^ In a subset of records, we measured the inter-assessor agreement. Both assessors were blinded for relationships identified from the genealogical data and for each other’s assessments. Furthermore, for each patient, we assessed whether the *C9orf72* HRE was inherited paternally or maternally, based on clinical family history or inferred from reconstructed pedigrees.

### C9orf72 Haplotype Reconstruction and Phenotypic Heterogeneity

We reconstructed all haplotypes on chromosome 9 spanning the *C9orf72* HRE and identified shared IBD segments longer than 1 centimorgan (cM), consistent with a shared ancestor approximately 50 generations back. First, we lifted the GRAPE sequencing data over from GRCh37 to GRCh38 and harmonized to a TOPMed reference panel.^26^ A surrogate marker for the *C9orf72* HRE was inserted at chromosome 9 position 27,573,534 (GRCh38). Chromosome 9 genotypes of 238 *C9orf72* HRE carriers were then phased using SHAPEIT5,^27^ jointly with 3583 Dutch reference samples from Project MinE, also harmonized to the TOPMed reference panel.

To validate phasing of the surrogate *C9orf72* HRE, we calculated Hamming distances between haplotypes of *C9orf72* carriers and the published^13,28,29^ European *C9orf72* founder haplotype. Phasing was considered correct when the surrogate HRE marker was located on the haplotype showing more than 90% concordance with the published founder haplotype. When the marker was placed on the discordant allele, *C9orf72* HRE phasing was shifted to the other allele. If both alleles were highly concordant with the European haplotype, phasing was accepted without modification. When both alleles were discordant, we visually inspected the local haplotype structure around *C9orf72* to assign the *C9orf72* HRE–carrying allele.

Then, we performed two separate IBD detection analyses. First, we detected shared IBD segments specifically around the *C9orf72* locus. We filtered for IBD segments surrounding the *C9orf72* HRE with a minimum IBD segment length greater than 1 centimorgan (cM), corresponding approximately to a shared ancestor up to 50 generations ago. For this analysis, we removed the non-*C9orf72* haplotype in *C9orf72*-carrying individuals. Second, we ran a genome-wide IBD detection analysis among *C9orf72* carriers and computed how many cM of IBD segments they shared. For both IBD analyses we used HapIBD with endpoint refinement, with HapIBD parameters set lenient as not to miss potential IBD segments: min-seed 0.5, max-gap 15000, min-extend 0.1, min-output 1.0, min- markers 50, min-mac 1.^30,31^ The 3583 Dutch Project MinE samples were initially included in the HapIBD analysis to improve the accuracy of endpoint refinement. Segments shared with these non- *C9orf72* reference samples were removed afterwards.

We then sought to address whether regional genetic variation at the *C9orf72* locus or at a genome- wide scale explains phenotypic heterogeneity in ALS caused by the *C9orf72* HRE. To this end, we regressed (regional) IBD sharing on age at onset and survival.

### IBD-Based Localization of the C9orf72 HRE

Genome-wide IBD segments from patients with a *C9orf72* HRE, generated as described above, were used to evaluate the precision and recall of IBD-based detection of the pathogenic *C9orf72* locus in distantly related ALS patients. Segment endpoints flanking the *C9orf72* locus were visually inspected, and false-positive or false-negative endpoints were corrected when appropriate. Within each extended pedigree, we identified IBD segments shared by all affected individuals across a range of minimum segment-length thresholds (1–10 cM). The size of each pedigree was measured by counting the number of unique meioses connecting all affected individuals to their most recent common ancestor. We defined precision as the total genomic length in cM of IBD segments shared by all patients in the pedigree, and recall as the proportion of pedigrees in which the shared segments encompassed the *C9orf72* locus. Precision and recall were evaluated across segment- length thresholds and meiosis counts.

### Statistical Analyses

Statistical analyses were performed using R 4.5.3.^32^ We used the ggplot2, pheatmap, and pedtools^33^ packages to generate plots and visualize pedigree structures. Correlations were quantified using both Pearson’s and Spearman’s rank correlation coefficients, and corresponding p-values were reported. Statistical significance was evaluated at a threshold of P < 0.05 and interpreted alongside effect sizes.

## Results

### Extended pedigree reconstruction

We obtained genealogical and clinical data for 238 ALS patients carrying a *C9orf72* HRE. Baseline demographic and clinical characteristics of the cohort are summarized in Table 1. The ancestor tables from the NRO archives included 3332 ancestors, dating back to 1810 (Table S1). For 3218 individuals, the first and last name were available, which is the minimum requirement for ancestor matching.

**Table 1.** Characteristics of *C9orf72* repeat expansion carriers. Survival was defined as time from symptom onset to death; PMA indicates progressive muscular atrophy. Data are presented as *n* (%) for categorical variables and median (IQR) for continuous variables.

| **Variable** | **Value** |
|---|---|
| **Sex** |  |
| Female | 114 (48%) |
| Male | 124 (52%) |
| **Age at onset, years** | 61 (55-67) |
| **Diagnosis** |  |
| ALS | 222 (93%) |
| ALS-FTD | 11 (4.6%) |
| PMA | 5 (2.1%) |
| **Survival, months** | 20 (13-29) |
| **Family history** |  |
| fALS | 110 (46%) |
| Suspected fALS | 11 (4.6%) |
| sALS | 117 (49%) |

Using an automated custom pipeline (Figure 1), we detected shared ancestors and reconstructed extended pedigrees. After ancestor matching and additional querying of public CBG genealogy archives, 3228 unique individuals (index patients and ancestors) remained in the dataset, dating back to 1798.

Based on the ancestor table data alone, 71 patients were included in one of 31 extended pedigrees, yielding 49 pairs of related patients up to the sixth degree of relatedness (second cousins once removed). Incorporating genetic relatedness and subsequently retrieving ancestors from public archives, we identified a total of 91 probands across 39 extended pedigrees, revealing 67 patient pairs related up to the eighth degree (third cousins once removed). These pedigrees connected (apparently) sporadic patients to known families (25 pairs, 37%), to other (apparently) sporadic patients (19 pairs, 28%), or linked multiple known families (20 pairs, 30%, Figure 2).

**Figure 2:**
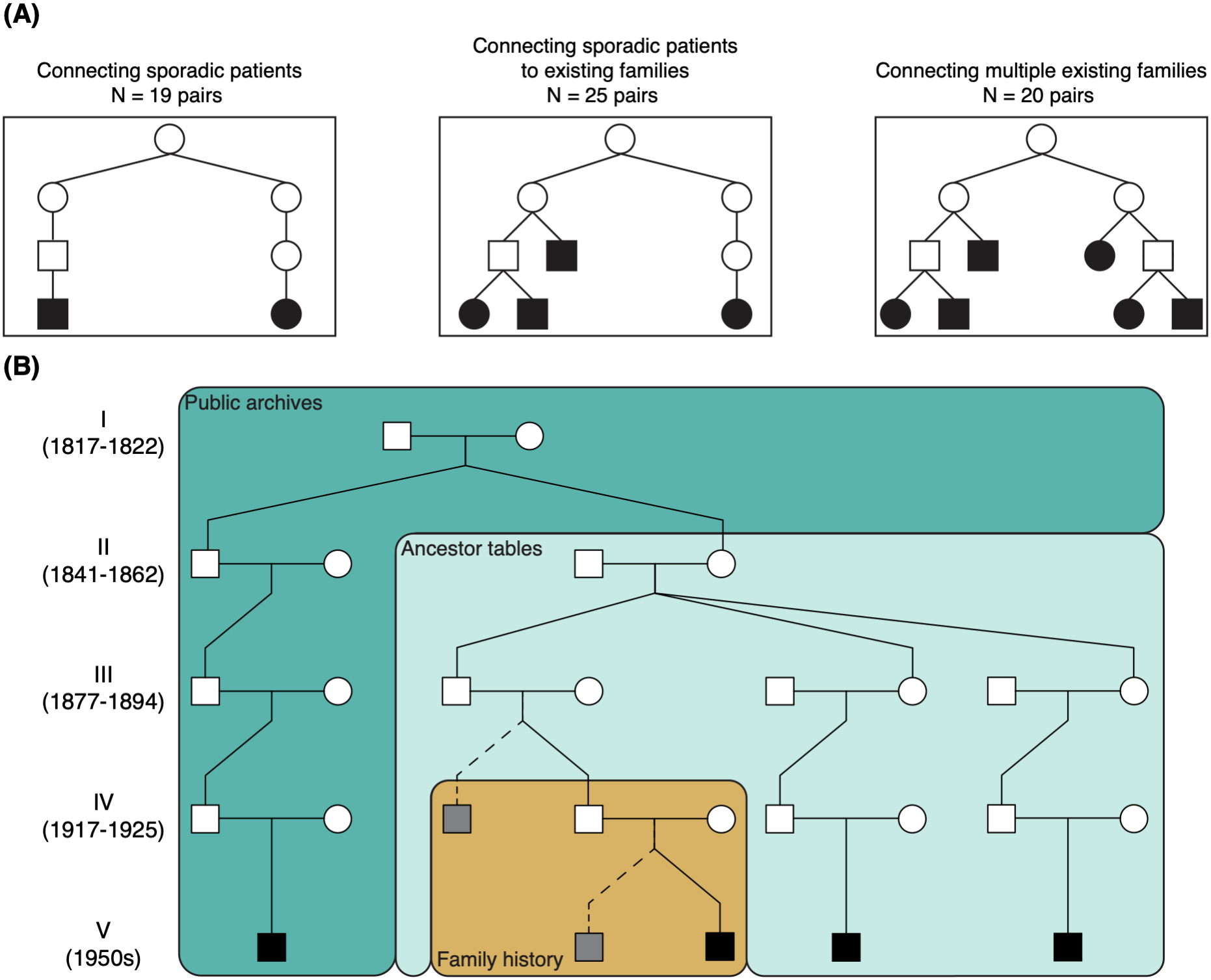
Reconstructed extended pedigrees. (A) Overview of types of extended pedigrees constructed from the archive data. (B) Example pedigree, colored by the reach of the different data sources. The range of birth years for each generation are indicated. Note that the individuals in gray with dashed lines are not part of the cohort analyzed in this study, but are added only for illustrative purposes.

Next, we compared archive-based relatedness with relatedness estimates from the GRAPE pipeline^21^ (Figure 3A). Among the 238 patients, GRAPE detected 77 pairs related in the seventh degree or closer. Using only the ancestor table data, which generally extends to the sixth degree, 49 pairs (64%) were confirmed within 1 degree of relatedness. When public archive data were included, 64 pairs (83%) were confirmed within 1 degree, and 2 pairs (3%) within 2 degrees. The additional public archive data also revealed 1 eighth-degree pair that GRAPE missed. The remaining 11 pairs (14%) could not be verified, but in many cases probands had ancestors from the same area in the Netherlands. This can cause a high level of background genetic relatedness in case of an isolated population. Alternatively, unregistered ancestral events, such as a child born out of wedlock, will result in genetic relatedness that cannot be confirmed in the official archive records.

**Figure 3:**
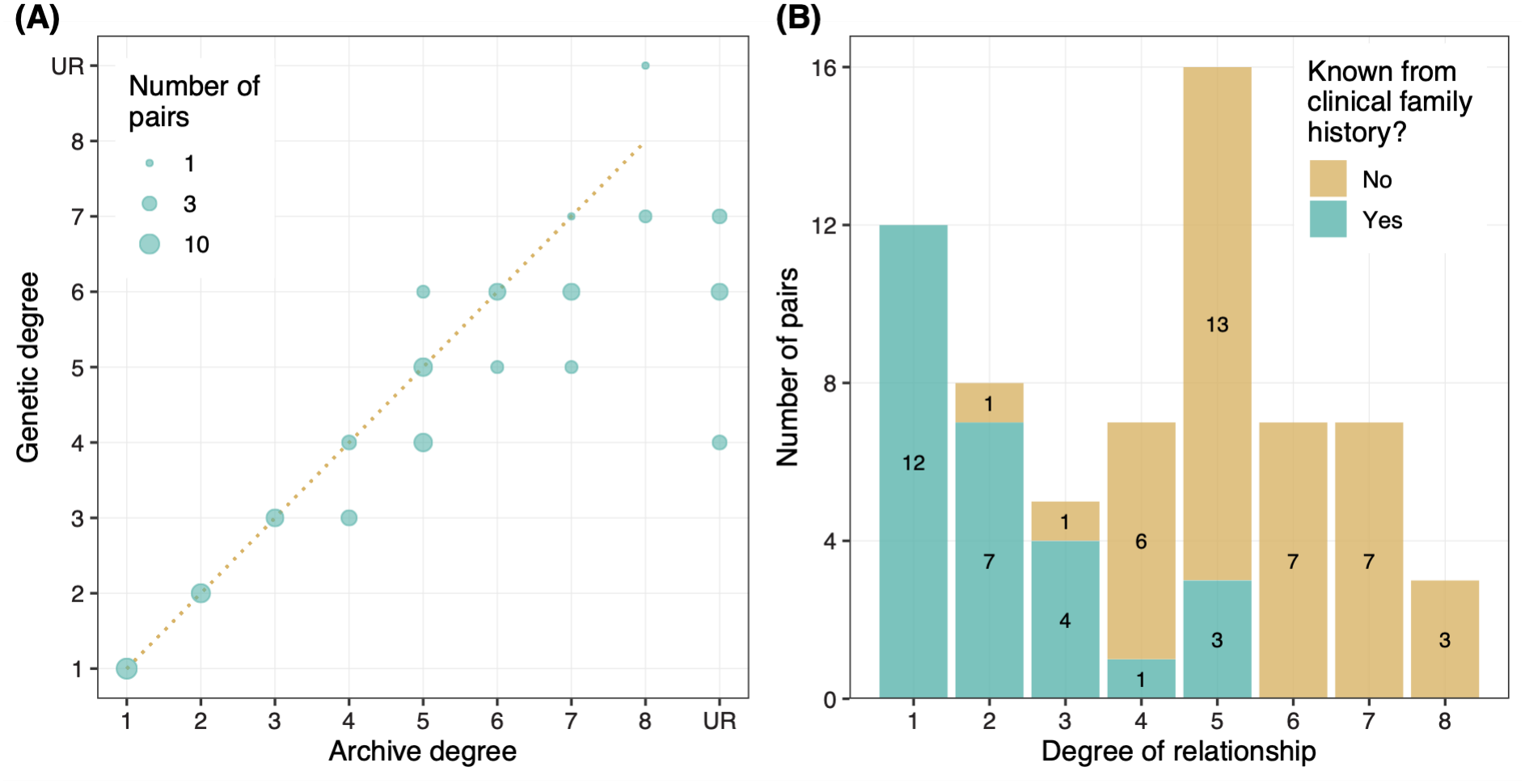
Accuracy and power of extended pedigrees. (A) Comparison of relationships found with archive and genetic data. (B) Overview of relationships that could and could not have been identified from clinical family history assessment, split by degree of relatedness.

For patients whose clinical family history was reevaluated, agreement between the two assessors was high (Cohen’s kappa 0.87). Despite this, 53% of patients with an affected relative identified in the extended pedigrees reconstructed from archive records were labeled as sporadic based on the clinical family history (following the definition of fALS as at least one first- or second-degree relative with ALS or FTD). Most relationships up to the third degree (23/25) could have been identified based on clinical records alone (Figure 3B). For relatives more distant than third degree (first cousins), only 10% (4/40) of relationships were documented in clinical records. Assessors also scored the putative parental transmission (maternal or paternal), which is clinically used to inform genetic counseling.

For 27 patients, the parent who transmitted the *C9orf72* HRE could be inferred from both clinical family history and the reconstructed extended pedigree. In 3 patients (11%), the parental origin of the *C9orf72* HRE was incorrectly inferred from the clinical family history. In these cases, reported non-specified dementia in a relative was assumed to be *C9orf72* HRE-related, leading to the incorrect parent being identified as the origin.

### C9orf72 haplotype and phenotypic correlates

In total, 234 patients carried the *C9orf72* HRE on the reported European founder haplotype. In 4 samples, both haplotypes were >10% discordant with the European founder haplotype. In 3 of these patients, the *C9orf72* HRE haplotype was shorter than the published European founder haplotype,^13,28,29^ likely due to recent recombination (Figure S2A, individuals 2-4). One patient carried the *C9orf72* HRE on a distinct haplotype (Figure S2A, individual 1). This individual was of European ancestry as inferred by principal components analysis (Figure S2B). We found no evidence of a second founder effect of the *C9orf72* HRE, as this distinct haplotype was not shared with others and could have been created by two recent recombination events. We found that the correlation between haplotype sharing around the *C9orf72* locus and genome-wide IBD (Figure S3) is highly significant but explains little variance (R^2^ = 0.215, p < 0.001).

Next, we assessed whether sharing genetic variation in the *C9orf72* haplotype could explain phenotypic similarities in patients with ALS caused by the *C9orf72* HRE (Figure S4). Shared IBD segment length at the *C9orf72* locus was not associated with phenotypic similarity in age at onset or the rate of disease progression measured as survival after disease onset. Similarly, we found no correlation between the amount of genome-wide IBD sharing and phenotypic similarity in age at onset or survival.

### Power of Extended Pedigrees for Disease Locus Mapping

To assess the power of extended pedigrees to map disease loci, we included all pedigrees with at least 2 meioses linking all patients to a common ancestor. In total, 89 patients were connected in 38 extended pedigrees (range: 2-4 patients; 2-14 meioses, Figure 4A-B). Pedigrees with more than 2 patients were additionally subdivided into smaller pedigrees, yielding 87 unique pedigree structures in total (Figure 4B). In each pedigree, we assessed the number of shared IBD segments between all patients (Figure 4C).

**Figure 4:**
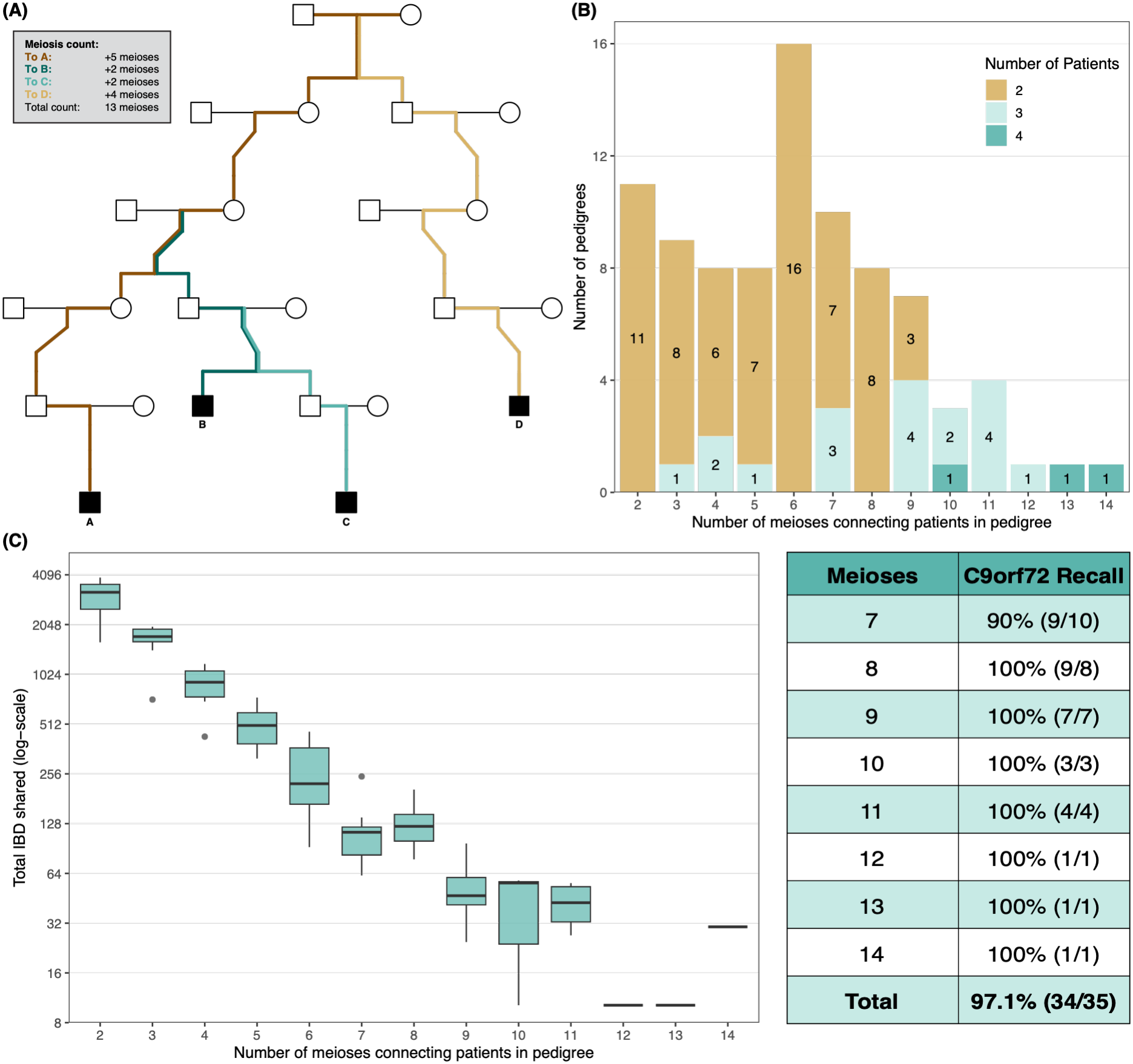
IBD-based detection of pathogenic C9orf72 locus in constructed extended pedigrees. (A) Meioses count. The complexity of each pedigree is computed as the number of unique meioses required to connect all affected individuals in the pedigree. (B) Number of extended pedigrees connected by number of meioses. The number of affected individuals per pedigree is displayed in different colors. (C) Precision and recall of IBD-based analysis in detecting the pathogenic *C9orf72* locus when considering IBD segments >4 cM long. Precision is defined as the total genetic length in cM that is IBD among all affected individuals in a pedigree. Boxplots illustrate median and IQR. Recall is defined as the proportion of pedigrees in which the *C9orf72* locus was captured in IBD segments.

Precision and recall of shared IBD segment detection, as a function of pedigree meiosis count and minimum shared segment length, are shown in Figure S5. Precision was poor in pedigrees connected by 6 or fewer meioses (n = 52), as on average ≥202.0 cM was shared between all patients. Similarly, minimum segment lengths greater than 1 cM yielded poor precision owing to numerous noncausal shared segments from more distant common ancestors, or technical IBD-detection artifacts. In contrast, for 34 out of 35 (97%) pedigrees with 7 or more meioses (n = 35), the *C9orf72* HRE was identified on shared segments longer than 4 cM, within a mean of 10.2-127.0 cM of total shared IBD. This corresponds to 0.13-1.72% of the entire genome. In the largest three pedigrees of 12 to 14 meioses, only 10 to 30 cM of IBD segments were shared. Notably, in a four- or five-generation pedigree, 3 or 4 affected individuals were typically required to reach 7 meioses (Figure 4B).

## Discussion

Genetic analyses in pedigrees remain a powerful approach to identify rare disease-causing variants, but finding large enough pedigrees can be difficult in a late-onset disease such as ALS. Using nationwide genealogical archives, we reconstructed extended pedigrees for 238 ALS patients carrying the *C9orf72* repeat expansion. In total, 91 patients were members of 39 multigenerational pedigrees with relationships up to the eighth degree dating back to ∼1800. Of these, 48 (53%) were clinically classified as sporadic. The extended pedigrees substantially exceeded what was captured by clinical family history alone, in which relationships beyond the third degree were rarely recognized.

Furthermore, in 11% of patients, the parental origin of the *C9orf72* HRE in the family had been incorrectly inferred from the clinical family history alone, which can hinder adequate genetic counseling. Taken together, this illustrates that leveraging archive records overcomes the limitation of clinical family histories which are often insufficient to ascertain the underlying genetic contributions to disease.

Compared with genetic relatedness inference, archive-based relationships agreed within 1 degree for more than 80% of pairs, supporting the accuracy of genealogical reconstruction. Additionally, genetic relationships provided clear directions for further archive research leading to the identification of more related pairs. Both approaches have complementary strengths. Genealogy-based pedigrees provide reliable, structured family relationships that are directly amenable to linkage and segregation analyses. Genetic relatedness estimates, on the other hand, can verify biological relatedness and identify more distant relationships. Although most genetic methods do not reconstruct pedigree structures,^21,24,34–36^ inferred relatedness can serve as input for targeted genealogical analyses.

Reconstruction of extended pedigrees is a powerful approach to map causal genes. The search space for a causal mutation decreases exponentially with the number of meioses separating patients in a pedigree. As a result, in pedigrees where patients are separated by at least 7 meioses, shared IBD segments longer than 4 cM consistently identified the *C9orf72* repeat expansion within a limited set of IBD segments spanning at most 127.0 cM. This further decreased with each additional meiosis, down to 10.2 cM in the largest pedigrees. These results indicate that IBD-based approaches in extended pedigrees can help identify pathogenic variants in ALS.

We did not observe a correlation between the amount of IBD sharing on the *C9orf72* haplotype and phenotypic similarities, such as age at onset or survival, among patients with the *C9orf72* repeat expansion. This could indicate that remote genetic variation outside the shared C9orf72 locus, or repeat instability between generations modifies these clinical characteristics.^29^ This motivates in-dept analyses of the *C9orf72* HRE itself or genome-wide screens for modifiers within these pedigrees, an approach similar to studies that found genetic modifiers of age at onset in Huntington’s disease.^37,38^

Our framework is generalizable and can be applied to any disease with a similar genetic architecture to ALS or heritable diseases that are often underreported in family histories or medical records.^5^ While the Mangrove pipeline is designed to work with archive data, other sources of pedigree data can easily be imported as well. For example, as shown in this study, one can include additional archive records obtained through a targeted search informed by inferred genetic relationships.

Furthermore, pedigree data from family histories obtained in the clinic or a research setting may be incorporated.

Some limitations merit consideration. First, the size of reconstructed extended pedigrees was constrained by the depth of genealogical data available from the archive records. These currently extend approximately 3 generations and thus allow reliable detection of relatives up to the sixth degree. Combined with inferred genetic relatedness and records obtained from public archives, however, related pairs were found up to the eighth degree. This was sufficient to narrow down the search space for a pathogenic variant to <2% of the entire genome using haplotype analysis in pedigrees including 7 meioses. Second, this framework relies on suitable archive records, which might not always be available or accessible in every country. With large-scale efforts to digitalize archives such as the eArchiving Initiative Consortium, however, these data will become increasingly available to clinicians, researchers and the general public.

Finally, as more distant relatives are considered, not all related patients might share the same monogenic cause of ALS. In these so-called phenocopies, ALS can occur due to polygenic risk or a different pathogenic mutation inherited from a distinct founder (see example in Figure S6). In these families, searching for a shared haplotype among all affected members of the pedigree may fail to identify a single shared causal variant. This can be mitigated in large pedigrees by subgrouping patients in different branches of the pedigree.

This study demonstrates that integrating extended genealogy with IBD analysis provides a scalable and powerful framework for detecting shared pathogenic loci among distantly related ALS patients. This approach can be extended to other late-onset heritable diseases where clinical family histories are often incomplete. This strategy can be applied in larger cohorts for gene discovery and variant interpretation in the clinic.

## Data and code availability

There are restrictions to the availability of the data due to its privacy-sensitive nature. The code for processing the archive data and reconstructing extended pedigrees, the Mangrove software toolkit, is freely available on GitHub (https://github.com/dvanoosten/mangrove)

## Data Availability

There are restrictions to the availability of the data due to its privacy-sensitive nature. The code for processing the archive data and reconstructing extended pedigrees, the Mangrove software toolkit, is freely available on GitHub.

https://github.com/dvanoosten/mangrove

## Acknowledgements

We thank Roy Verhoeven, Pieter van de Polder, Martine Zoeteman-van Pelt, Dorien van Driel and Roosje Keijser at the Dutch Center for Family History for providing the archive data.

This work was supported by the Dutch ALS Foundation (GoALS project, AV2021-002) and the Debman Foundation. W.v.R. was supported by the Frank’s PEDALS fellowship. M.A.v.E. has received funding support from the Netherlands Organisation for Health Research and Development (Vidi scheme). This work was sponsored by the Dutch Research Council (NWO–Domain Science) for the use of supercomputer facilities.

## Author contributions

D.v.O., P.B., and B.W. contributed equally. J.H.V. and W.v.R. jointly supervised this work. Conceptualization: J.H.V. and W.v.R., Data Curation: D.v.O., P.B., B.W., S.J.P., N.W., K.v.d.B., M.P.T.B., M.M., N.D.v.d.S., H.V.-B., A.R.K., L.v.d.V., M.A.v.E., L.H.v.d.B., J.H.V., and W.v.R., Formal Analysis: D.v.O., P.B., and B.W., Methodology: D.v.O., P.B., B.W., J.H.V. and W.v.R., Writing – original draft: D.v.O., P.B., B.W., J.H.V, and W.v.R., Writing – review and editing: S.J.P., N.W., K.v.d.B., M.P.T.B., M.M., N.D.v.d.S., H.V.-B., A.R.K., L.v.d.V., M.A.v.E., L.H.v.d.B., Funding Acquisition: L.H.v.d.B., J.H.V., and W.v.R., Supervision: J.H.V. and W.v.R.

