## Supplemental information for "A Framework For Reconstruction And Analysis Of Extended Pedigrees From Large-Scale Genealogy And Archive Records"

| **Generation** | **Total number**  **of individuals** | **First name available** | **Last name available** | **Place of birth available** | **Date of birth available** | **Date of death**  **available** | **Date of birth (median, range)** |
| --- | --- | --- | --- | --- | --- | --- | --- |
| Parents | 476 | 475 | 475 | 469 | 469 | 440 | 1919 (1888-1950) |
| Grandparents | 952 | 927 | 926 | 911 | 909 | 886 | 1888 (1843-1932) |
| Great-grandparents | 1904 | 1819 | 1817 | 1152 | 1120 | 857 | 1862 (1810-1909) |

### Table S1. Completeness of ancestor tables.

For each generation of ancestors, the (theoretical) total number of individuals is indicated, as well as the number of individuals with available data for several personal details. For date of birth, median and range are shown. Note that this has been calculated based on the available dates of birth only.

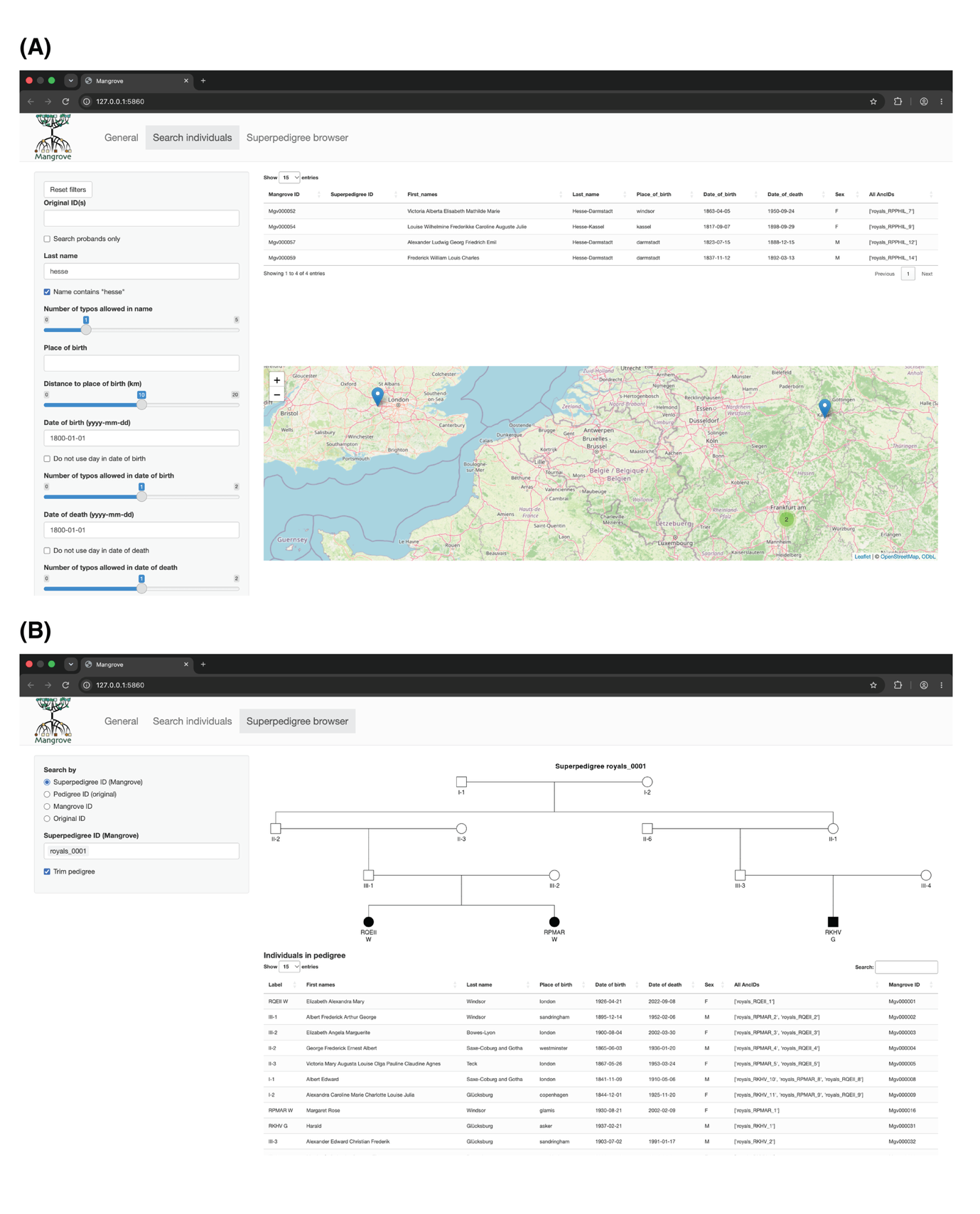

### Figure S1. Mangrove databrowser.

**(A)** Individual search tab, here searching for royal ancestors whose last name contains ‘hesse’. **(B)** Pedigree browser tab, here showing the pedigree of Queen Elizabeth II and Princess Margaret, and King Harald V.

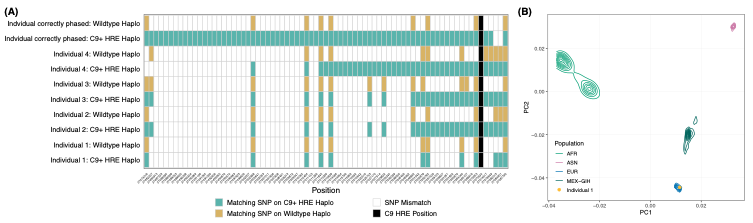

### Figure S2. Comparison of phased Dutch *C9orf72* haplotypes to published European *C9orf72* founder haplotype.

**(A)** Individuals 1-4 had 2 haplotypes on which the C9+ and wildtype haplotypes after phasing were discordant (>10% mismatch) with European founder haplotypes. The top individual shows correct phasing. **(B)** PCA projection of individual 1 on HapMap3 reference panel confirms European ancestry.

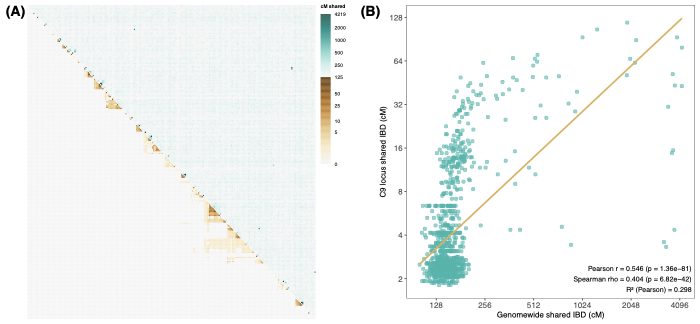

### Figure S3. Correlation of shared IBD segment around *C9orf72* HRE locus and total genome-wide shared IBD segments.

**(A)** Clustered heatmap of shared inter-individual IBD length around *C9orf72* HRE (brown, lower triangle) and total genome-wide IBD sharing (blue, upper triangle). Clustering was based on IBD sharing at the *C9orf72* HRE locus. **(B)** Scatterplot of IBD length around *C9orf72* HRE vs. genome-wide IBD sharing. Data were log-transformed as IBD sharing halves with every meiosis.

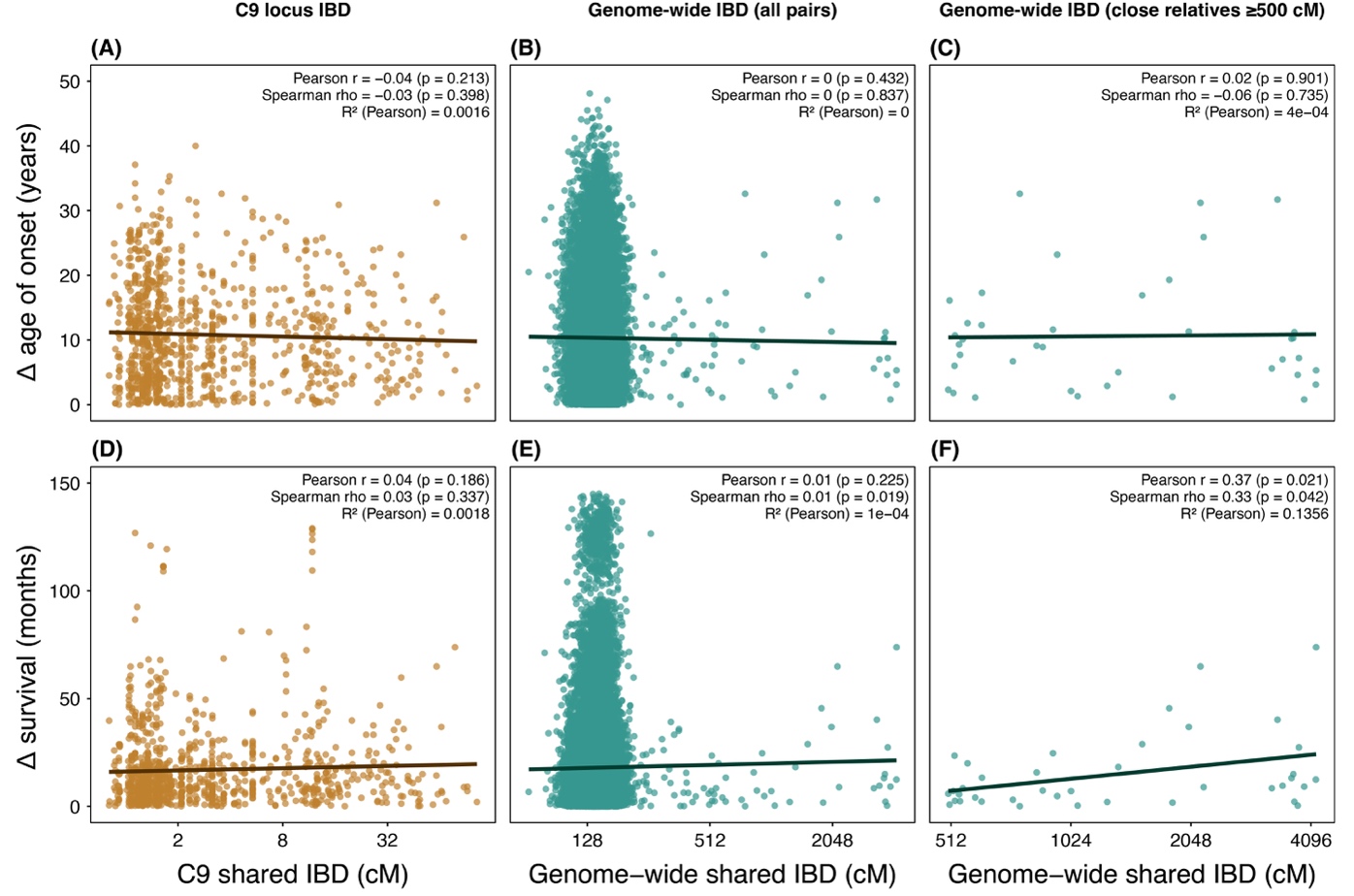

### Figure S4. Relationship between shared IBD and pairwise differences in clinical variables.

Pairwise differences in (**A–C**) age of onset and (**D–F**) survival are plotted against shared IBD length at the C9 locus (**left**), genome-wide across all pairs (**middle**), and among close relatives (**right**; IBD ≥ 500 cM). IBD lengths are shown on a log2 scale. Each point represents a pair of individuals; lines indicate linear regression fits. Pearson (r) and Spearman (ρ) correlation coefficients, corresponding p-values, and the coefficient of determination (R2) are reported.

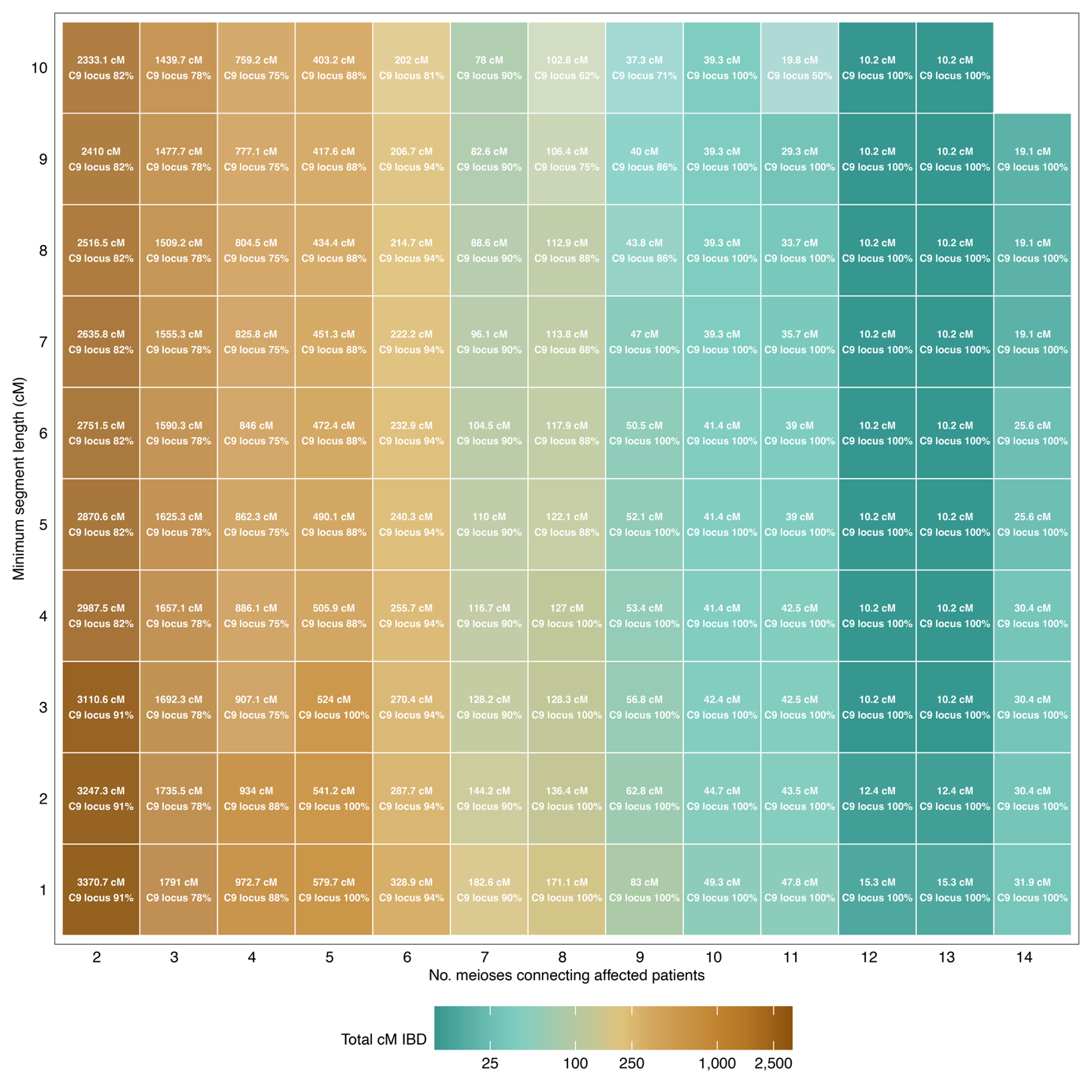

### Figure S5. Recall and precision of pathogenic *C9orf72* HRE detection.

Heatmap illustrating the percentage of pedigrees in which the pathogenic *C9orf72* HRE was among detected IBD segments shared by all affected individuals in the pedigree (recall) and the total genetic length of shared-by-all IBD segments (precision).

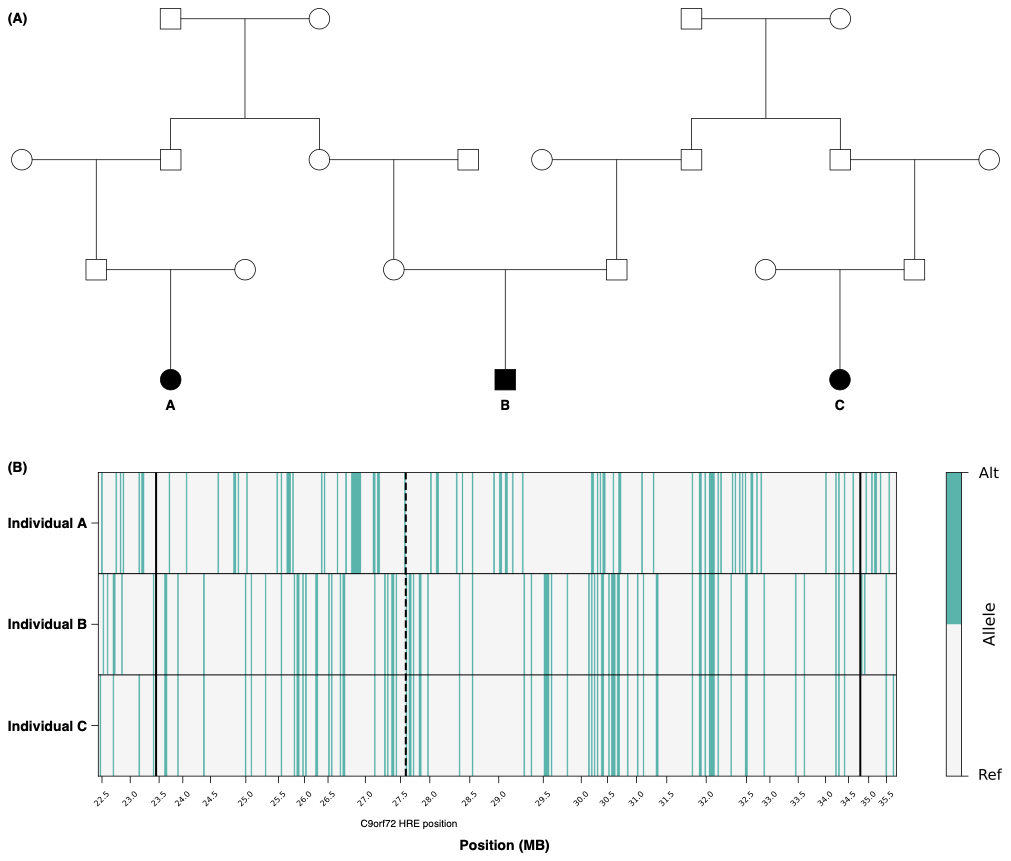

### Figure S6. *C9orf72* extended pedigree with multiple founder effects.

**(A)** Reconstructed extended pedigree of *C9orf72* HRE carriers showing that the *C9orf72* HRE in individual A did not originate from the same most recent common ancestor as those in individuals B and C. **(B)** Visualization of the shared IBD segment between individuals B and C. The black dashed line indicates the position of the *C9orf72* HRE (chr9:27573534, GRCh38). The black solid lines indicate the start and end of the shared IBD segment, resulting from HapIBD.
